# Does contrast-enhancement improve visualisation of lenticulostriate arteries in cerebral small vessel disease using time-of-flight magnetic resonance angiography at 7 Tesla?

**DOI:** 10.1101/2022.03.23.22272852

**Authors:** Christopher N. Osuafor, Catarina Rua, Andrew D. Mackinnon, Marco Egle, Philip Benjamin, Daniel J. Tozer, Christopher T. Rodgers, Hugh S. Markus

## Abstract

**Background And Purpose:** 7 Tesla-field-strength (7T) Magnetic Resonance Imaging (MRI) allows the small perforating arteries in the brain to be visualised, and this modality may allow visualisation of the arterial pathology in cerebral small vessel disease (cSVD). Most studies have used standard Time-of-Flight (ToF) Magnetic Resonance Angiography (MRA). Whether the use of contrast enhancement improves perforating artery visualisation at 7T remains unclear.

**Materials And Methods:** In a prospective study, we compared standard ToF MRA with contrast-enhanced (CE) ToF MRA at 7T for the visualisation of the lenticulostriate arteries (LSAs). Ten patients with symptomatic lacunar stroke were recruited (mean age, SD, 64±9.9 years). Visualisation was assessed using a visual rating scale administered by two independent expert readers and length of the LSAs visible.

**Results:** Visualisation of the LSAs was improved with CE ToF MRA. The mean Visibility and Sharpness Score was higher for CE ToF MRA over standard ToF MRA (2.55±0.64 vs 1.75±0.68; *P*=0.0008). The mean length of LSA visualised was significantly longer with CE ToF MRA compared to standard ToF MRA (24.4±4.5 vs 21.9±4.0 mm; *P*=0.01).

**Conclusion:** CE ToF MRA offers improved visualisation of the LSAs over standard ToF MRA. The addition of contrast may improve the ability to visualise cerebral small vessel disease arterial pathology.

## Introduction

Disease of the small cerebral perforating arteries, which supply the white matter and deep grey matter structures (cerebral small vessel disease, cSVD), causes a quarter of all strokes (lacunar strokes) and is the most common pathology underlying vascular cognitive impairment.^1^ Until recently, it has not been possible to visualise the perforating arteries in humans using non-invasive techniques. More recently, high resolution 7 Tesla-field-strength (7T) Magnetic Resonance Imaging (MRI) has allowed visualisation of perforating arteries because of the intrinsic increase in signal-to-noise ratio compared to lower field strengths at 1.5T or 3T.^2–4^ Studies have reported diffuse abnormalities in the perforating arteries in individuals with both stroke^5^ and stroke risk factors such as hypertension^6^, and have identified disruption in flow in individual perforating arteries associated with lacunar infarcts. 7T MRI has the potential to provide important insights into the arterial pathology associated with lacunar stroke and cSVD.

Many studies using 7T MRI to date have used standard Time-of-Flight (ToF) Magnetic Resonance Angiography (MRA) without contrast.^3–6^ Contrast-enhanced (CE) ToF MRA is the collective name for several pulse sequences that acquire rapid sequential 3-dimensional (3D) imaging following injection of a bolus of a gadolinium-containing contrast agent.^7^ Whether the addition of a gadolinium based contrast agent improves visualisation of the small perforating arteries, typically between 200-500 μm in diameter, is unclear. Arterial blood affected by contrast agent, will have shorter T_1_, thereby yielding higher signal than tissues not affected by the contrast agent.^8^ CE ToF MRA is widely used for visualisation of the larger arteries, including the extracerebral carotid and vertebral arteries for the visualisation of stenosis.^9,10^ Studies have suggested that contrast administration provides increased signal to noise and improved visualisation compared to standard ToF MRA,^2,11^ but it could also compromise visualisation by improving the visibility of veins, thereby increasing venous contamination. One 7T study retrospectively analysed data from a mixed cohort of patients, and suggested contrast enhancement may increase the length of perforating arteries which can be visualised,^12^ but no systematic analysis of its benefits has been performed.

In this prospective study of patients with lacunar stroke secondary to cSVD, we compared standard ToF MRA with CE ToF MRA at 7T. We assessed: 1) whether there was any improvement in radiological visualisation of the lenticulostriate arteries (LSAs); 2) the length of the individual LSAs that could be identified, and 3) the signal within the LSAs to noise ratio. We hypothesized that CE ToF MRA improves the delineation of LSAs compared to standard ToF MRA at 7T. To determine its utility in patients with cSVD, in whom perforating artery visualisation may be inferior to normal controls, we performed the comparison in individuals with symptomatic lacunar strokes.

## Materials and Methods

### Study Population

Inclusion criteria was a clinical lacunar syndrome with an anatomically corresponding lacunar infarct on brain MRI. Participants with any other potential cause of stroke other than cSVD (including large artery disease causing a stenosis > 50%, or a cardio-embolic source), or with any other potential cause of white matter disease (such as multiple sclerosis) were excluded. Additionally, subjects with a contraindication for 7T MRI (for example, claustrophobia, specific metal objects in or on the body, pregnancy, known allergy to gadolinium-containing contrast agent, or impaired renal function with estimated glomerular filtration rate of less than 59 mL/min/1.73m^2^) were excluded. This study was approved by the Institutional Review Board of East of England - Cambridge Central Research Ethics Committee (REC Ref: 19/EE/0219). Written informed consent was obtained from all participants. Participants were recruited between February 2020 and November 2020.

### MR Imaging Protocol

A 3D ToF MRA technique was performed on a whole-body human 7T MR system (7T Terra, Siemens Healthineers, Erlangen, Germany) equipped with a 32-channel receive coil (Nova Medical, Wilmington, Massachusetts). Before image acquisition, a 20-gauge intravenous cannula was placed in the antecubital vein for contrast injection. Firstly, ToF MRA pulse sequences were acquired; this was followed by manual intravenous administration of 0.1 mmol/kg of a gadolinium-based contrast agent [Gadobutrol, Gadovist®, Bayer PLC, Reading, UK)^13^ and 10 millilitres of 0.9% sodium chloride flush and finally, acquisition of post contrast MRA pulse sequences. The CE ToF MRA pulse sequences were acquired two minutes after contrast injection in nine participants; in one participant, pulse sequences were acquired at the same time of contrast injection. No changes were made to the sequence parameters after contrast administration. The following protocol was used for both the pre and post contrast sequences: Field of view (FOV) 200 × 156.3 mm^2^, Voxel Size 0.24 × 0.24 × 0.32 mm^3^, repetition time (TR) of 13 ms, echo time (TE) of 5.1 ms and flip angle (FA) of 20 degrees. Two slabs were used with 80 slices per slab to shorten the time of flight of inflowing blood and, thus increase the blood signal. To accelerate image acquisition, a partially parallel acquisition mode, GeneRalized Autocalibrating Partially Parallel Acquisition (GRAPPA) with an acceleration factor of two was used. Acquisition time (TA) was 9 minutes and 53 seconds. Standard pulse sequences which include T_1_-weighted sequence, T_2_-weighted sequence, T_2_-weighted fluid-attenuated inversion recovery (FLAIR) and diffusion-weighted sequences, were also obtained for the purpose of identifying radiological markers of cSVD like number of lacunes/lacunar infarcts, grade of white matter hyperintensities, number of microbleeds and any abnormal brain lesions.

### Postprocessing and data analysis

All images were exported to an offline station equipped with Syngo® (Siemens) and Weasis® medical image viewer software for image processing. Multiplanar Reconstructions (MPR) were performed in coronal, sagittal and axial planes. Maximum Intensity Projections (MIPs) were reconstructed in the coronal and axial (slab thickness 15 mm) directions. The MIPs were focussed on the main trunk of the middle cerebral artery (MCA) and anterior cerebral artery (ACA) to centre the LSAs. 3D-volume rendered images were used to ensure anatomical landmarks of the MCA and ACA. LSAs arising from the MCA (lateral LSAs) and horizontal part of the ACA (medial LSAs) were identified. Standard pulse sequences were for used for standard reporting to assess for lacunar infarcts, white matter hyperintensities, enlarged perivascular spaces and microbleeds.

### Comparison of Standard ToF MRA and CE TOF MRA Images

#### a. Visual Rating Scale

A qualitative visual rating scale was used to compare standard ToF MRA with CE ToF MRA using a scale, modified from work done previously on vessel imaging.^14,15^ Rating was performed independently by two raters (one clinical research fellow, C.N.O. and one consultant neuroradiologist, A.D.M.) who were blinded to each other’s rating, with both images placed side by side. When scores differed significantly (>2 point difference) between the 2 raters, the images were re-evaluated together, and the score determined by consensus. Overall visibility and sharpness of the LSAs, presence of artefacts and extent of venous contamination were rated.

i. Visibility and sharpness of the LSAs was assessed on a four-point rating scale (0, no LSA visualized; 1, poorly defined LSAs; 2, well defined LSA; 3, excellent definition of the LSA). This was performed on the coronal MIPs.
ii. Presence of artefacts (subject-dependent motion artefacts and flow artefacts around the main trunk of the MCA and the ACA) was assessed on a four-point rating scale (0, no artefact affecting visibility; 1, mild artefacts; 2, moderate artefacts; 3, severe artefacts). This was performed on the original images before MIP reconstruction.
iii. Extent of venous contamination of the image was assessed as cerebral veins and the dural venous sinuses (as well as arteries) were enhanced on the CE images. It was determined whether enhancement of the venous system (especially the basal cerebral veins and internal cerebral veins and their tributaries) affected the ability of the rater to visualise the origin or course of the LSAs. This rating was done on the axial MIP based on a four-point rating scale (0, no venous contamination affecting visibility; 1, mild venous contamination; 2, moderate venous contamination; 3, significant venous contamination).

#### b. Length of LSA visualised

This was measured (in millimetres) as the length of the most lateral LSA as a straight (through-space) linear distance from the origin of the LSA at the MCA or ACA, to its most distal visible part. These measurements were performed by one rater on the coronal MIP and was reported for all subjects (Supplementary Fig 1). In addition, the number of visible LSAs originating from the MCA and the ACA was recorded.

#### c. Signal to Noise Ratio

The conspicuity of the LSA relative to the background was analysed by calculating the Signal to Noise Ratio (SNR) of the LSA, SNR of brain and Contrast to Noise Ratio (CNR) of the LSA to the background brain tissue. This was also done by a single rater. SNR was defined as 0.695 x (SI/σ),^16^ where SI is signal intensity of the blood in the LSA, σ is the standard deviation of the noise determined in a signal-free, artefact-free background region, and 0.695 is the Rayleigh distribution correction factor to calculate the true SNR. The SI of the blood in the LSA was measured on the most prominent LSA on either side, as close as possible to its origin (from the MCA or ACA) using a region of interest (ROI) area of 0.2mm^2^. SI of brain was measured using an ROI area of 20mm^2^ at an area of brain adjacent to the origin of the LSA. σ was measured on an ROI with area of 50mm^2^, drawn (in-air) at the posterior outer corner of the most inferior slice of the image. This was to avoid artefacts introduced into the background from the eyes (which are always in motion) in the anterior outer corners of the most superior slices. Finally, the CNR of the LSA to brain (CNR_LSA-BRAIN_) was defined as SNR_LSA_ minus SNR_BRAIN_,^16^ where SNR_LSA_ is the SNR of the LSA and SNR_BRAIN_ is SNR of brain. For SI measurement and placement of ROIs, an axial plane on the original data was used as this gave the best view of the LSAs; this also enabled drawing on the same plane, both the background noise standard deviation ROI and the brain ROI on the anterior (adjacent) aspect of the MCA (Supplementary Fig 2). When the LSAs were not visible on this plane, signal intensity was assessed on the coronal plane; in likewise fashion, the brain ROI was drawn adjacently.

### Statistical Analysis

Statistical analysis was performed using R statistical software version 3.4.1.^17^ Inter-rater variability between the two raters for the visual rating scale, was assessed with the Cohen’s weighted kappa using R package irr.^18^ The strength of the inter-rater agreement was categorized as follows: ≤ 0, no agreement; 0.01–0.20, none to slight; 0.21–0.40, fair; 0.41– 0.60, moderate; 0.61–0.80, substantial; 0.81–1.00 almost perfect agreement. The mean qualitative difference between standard ToF MRA and CE TOF MRA across the two raters was analysed with a Wilcoxon signed-rank test and plotted using R package ggpubr.^18^ The quantitative difference was also analysed using Wilcoxon signed-rank test for all measures except number of visible LSAs and Straight Length of LSAs where due to their parametric distribution, a paired t-test was employed. Statistical significance was defined as *P*<0.05.

## Results

### Subjects

Ten participants (four males, six females) were included. Mean age was 64±9.9 years (range 51-80 years). Demographic and clinical data of subjects are summarized in Table 1.

**Table 1.** Baseline demographics, vascular risk factors and radiological characteristics of participants.

| Characteristic | Patients (n=10) |
| --- | --- |
| Gender (Female) | 6 (60%) |
| Age at stroke onset, years | 64 (51–80) |
| Ethnicity (White British) | 9 (90%) |
| BMI, kg/m<sup>2</sup> * | 26.4 (25.6–30.8) |
| Modified Rankin Score at consent | 0 (0–2) |
| Vascular Risk Factors |  |
| Hypertension | 10 (100%) |
| Hyperlipidaemia | 9 (90%) |
| Diabetes Mellitus | 1 (10%) |
| Smoking History (current or previous) | 9 (90%) |
| Alcohol Intake ($\geq 14$ units/week) | 3 (30%) |
| Previous Stroke/TIA | 3 (30%) |
| Number of Lacunes/Lacunar infarcts |  |
| 1 | 5 (50%) |
| >1 | 5 (50%) |
| Fazekas Grading of White Matter Hyperintensities |  |
| Grade 0 | 0 |
| Grade 1 | 9 (90%) |
| Grade 2 | 0 |
| Grade 3 | 1 (10%) |
| Microbleeds |  |
| 0 | 9 (90%) |
| 1 | 1 (10%) |
| >1 | 0 |

Categorical variables are expressed in counts and percentages; ordinal and continuous variables are expressed in median (range), except for age which was expressed as mean (range). BMI, Body Mass Index; Fazekas grading of White Matter Hyperintensities at periventricular white matter 0 = absent; 1 = caps or pencil-thin lining; 2 = smooth halo; 3 = irregular periventricular signal extending into the deep white matter; and at deep white matter: 0 = absent; 1 = punctate foci; 2 = beginning confluence; 3 = large confluent areas. \*BMI not available in 2 patients.

### Visual rating scale

Inter-rater agreement for the qualitative analysis was moderate for Overall Visibility and Sharpness Score, fair to moderate for Presence of Artefact Score, and substantial to perfect for Extent of Venous Contamination Score (Table 2). Since inter-rater agreement between both raters was moderate to perfect indicating good agreement, the mean scores for both raters were used for the qualitative comparison of the standard ToF MRA with the CE TOF MRA. A summary of the qualitative analysis is shown in Fig 1. Overall Visibility sand Sharpness Score was higher for CE ToF MRA over standard ToF MRA (2.55±0.64 vs 1.75±0.68; *P*=0.0008).

**Table 2.** Inter-rater agreement between raters for qualitative analysis.

| Qualitative analysis scales | Pulse sequence | Percentage agreement between raters | Weighted Kappa | P value | Strength of agreement |
| --- | --- | --- | --- | --- | --- |
| Overall Visibility and Sharpness Score | Standard ToF MRA | 70% | 0.595 | 0.01* | Moderate |
|  | CE ToF MRA | 70% | 0.531 | 0.04* | Moderate |
| Presence of Artefacts Score | Standard ToF MRA | 60% | 0.375 | 0.09 | Fair |
|  | CE ToF MRA | 60% | 0.444 | 0.04* | Moderate |
| Extent of Venous Contamination Score | Standard ToF MRA | 100% | NA | NA | Perfect<sup>#</sup> |
|  | CE ToF MRA | 90% | 0.615 | 0.04* | Substantial |

Inter-rater agreement for the qualitative analysis was moderate for Overall Visibility and Sharpness Score, fair to moderate for Presence of Artefact Score, and substantial to perfect for Extent of Venous Contamination Score. NA = Not available as scores of zero (0) were not computed. <sup>#</sup>Perfect agreement based on 100% agreement between the two raters. Strength of agreement categorised as ≤ 0, no agreement; 0.01–0.20, none to slight; 0.21–0.40, fair; 0.41–0.60, moderate; 0.61–0.80, substantial; 0.81–1.00 almost perfect agreement. \*Statistical significance reached. ToF, Time-of-Flight; MRA, Magnetic Resonance Angiography; CE, Contrast-Enhanced.

**Fig 1.**
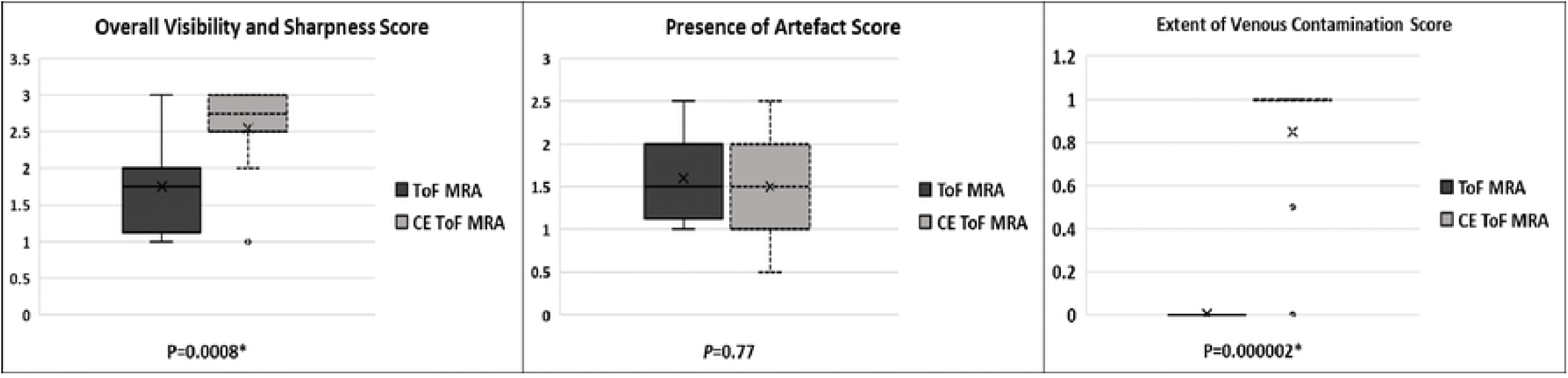
Qualitative analysis comparing standard ToF MRA and CE ToF MRA with boxplots and the Wilcoxon signed-rank test. For Overall Visibility and Sharpness Score, there was a statistically significant difference in favour of CE ToF MRA over standard ToF MRA. For Presence of Artefacts Score, there was no difference between standard ToF MRA and CE ToF MRA. For Extent of Venous Contamination Score, there was a difference in favour of standard ToF MRA over CE ToF MRA, however this was mild. **\***Statistical significance reached. ToF, Time-of-Flight; MRA, Magnetic Resonance Angiography; CE, Contrast-Enhanced.

An illustration of the improved visualisation seen with CE ToF MRA is shown in Fig 2.

For Presence of Artefacts Score, there was no difference (standard ToF MRA 1.6±0.52 vs CE ToF MRA 1.5±0.62; *P*=0.77). Venous Contamination Score was higher for CE ToF MRA over standard ToF MRA (1 vs 0; *P*=0.000002) as illustrated in Fig 3.

**Fig 2.**
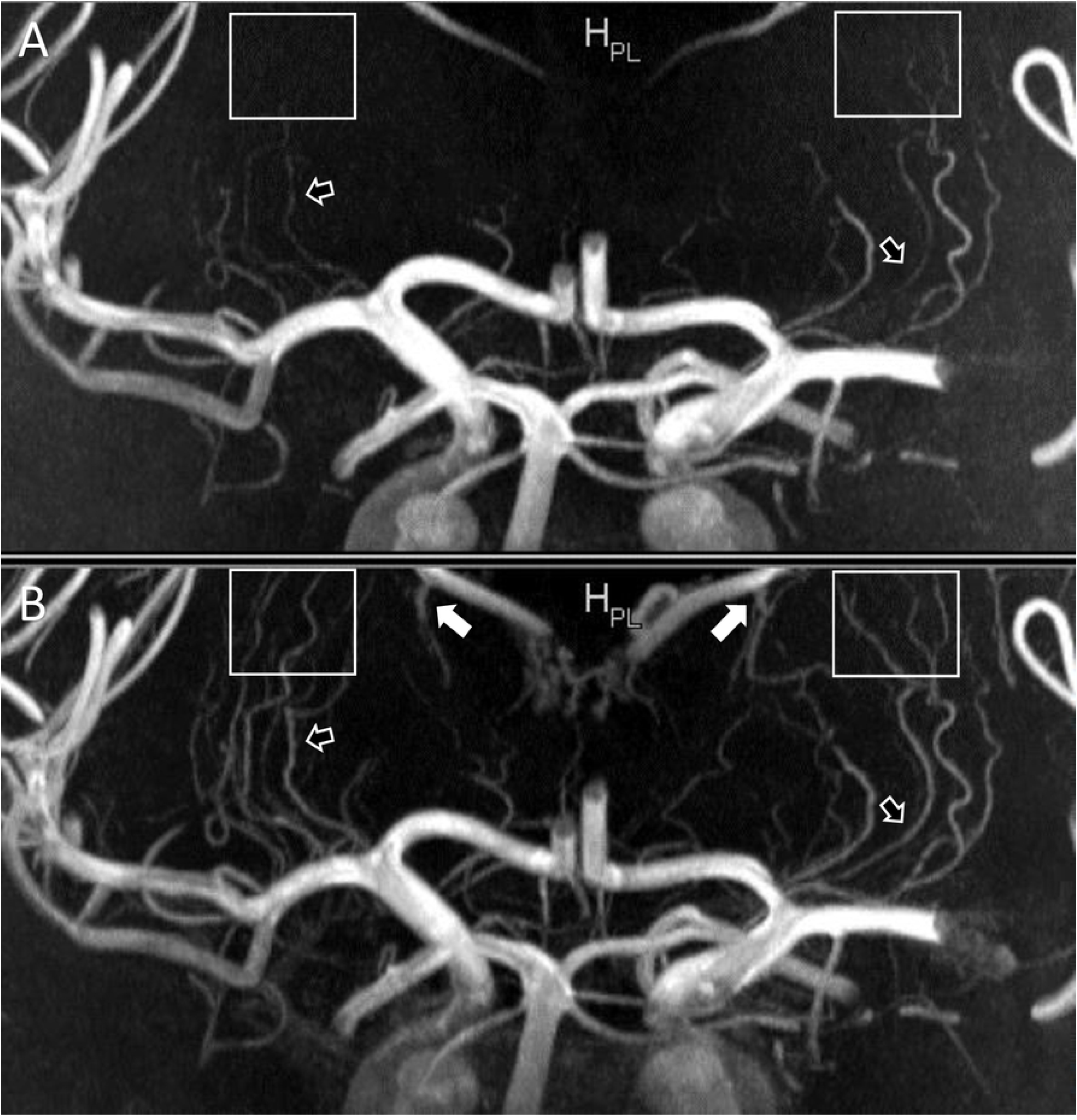
Coronal plane Maximum Intensity Projections comparing standard ToF MRA (A) and CE TOF MRA (B) for overall visibility and sharpness. The improved clarity of the LSAs in the CE ToF MRA and improved signal in areas of signal loss (black arrows with white borders) in the standard ToF MRA is shown. The LSAs could also be followed over a longer trajectory (white boxes). Also, prominence of the venous system (white arrows) is noted in the CE ToF MRA. For Overall Visibility and Sharpness Score scale, the standard ToF MRA was rated 2 (well-defined LSAs) while the CE ToF MRA was rated 3 (excellent definition of the LSAs). ToF, Time-of-Flight; MRA, Magnetic Resonance Angiography; CE, Contrast-Enhanced; LSAs, Lenticulostriate Arteries; MCA, Middle Cerebral Artery.

**Fig 3.**
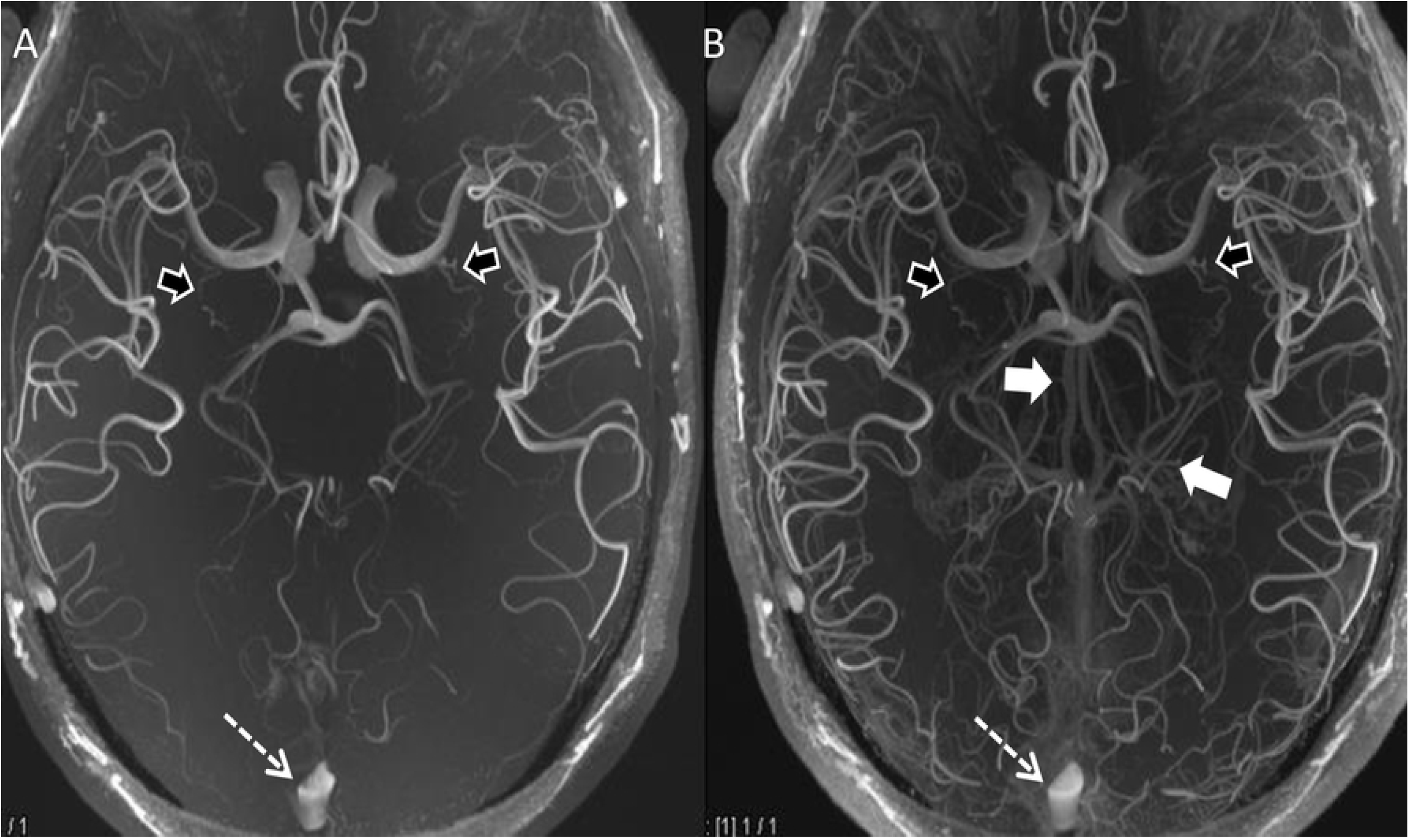
Axial plane Maximum Intensity Projections comparing standard ToF MRA (A) and CE ToF MRA (B) for extent of venous contamination. The prominence of the internal Cerebral Veins centrally and its tributaries (white arrows) and the Superior Sagittal Sinus (black arrows) which do not affect the visualisation of the LSAs at the MCA (white dashed arrows) is shown. For Extent of Venous Contamination Score scale, this was rated 0 (no venous contamination hampering visibility) in both images. ToF, Time-of-Flight; MRA, Magnetic Resonance Angiography; CE, Contrast-Enhanced; LSAs, Lenticulostriate Arteries; MCA, Middle Cerebral Artery.

### Length of LSA visualised

The mean length of LSA visualised was significantly longer with CE ToF MRA compared to standard ToF MRA (24.4±4.5 v 21.9±4.0 mm, *P*=0.01, Table 3). A figure format of the length measurements is shown in Supplementary Figure 2. There was no significant difference in the number of visible LSAs (4.6±2.1 vs 4.9±2.3, *P*=0.19, Table 3).

**Table 3:** Quantitative Analysis comparing standard ToF MRA, and CE TOF MRA based on Signal Intensities, Signal to Noise Ratio, Contrast to Nosie Ratio, number of visible LSAs and Straight Length of the LSAs.

| Quantitative analysis parameters | Standard ToF MRA | CE ToF MRA | *P* value |
| --- | --- | --- | --- |
| LSA Signal Intensity | 239.4±97.5 | 262.1±98.1 | 0.02* |
| Brain Signal intensity | 62.3±6.0 | 65.5±5.9 | 0.01* |
| Background Noise standard deviation | 4.3±0.4 | 4.5±0.5 | 0.08 |
| SNR<sub>LSA</sub> | 38.9±16.1 | 40.7±15.2 | 0.08 |
| SNR<sub>BRAIN</sub> | 10±0.5 | 10.1±0.7 | 0.77 |
| CNR<sub>LSA-BRAIN</sub> | 28.9±16.0 | 30.6±14.0 | 0.08 |
| Number of visualised LSAs | 4.6±2.1 | 4.9±2.3 | 0.19 |
| Straight Length of LSA (mm) | 21.9±4.0 | 24.4±4.5 | 0.01* |

All values expressed as mean ± SD (standard deviation). ToF, Time-of-Flight; MRA, Magnetic Resonance Angiography; CE, Contrast-Enhanced. LSA: Lenticulostriate Artery; SNR, Signal to Noise Ratio; CNR, Contrast to Noise Ratio; mm, millimeters. \*Statistical significance reached.

### Signal and Contrast to Noise ratios

The signal intensity of blood in the LSA was higher with CE ToF MRA compared to standard ToF MRA (*P*=0.02, Table 3). However, the brain signal intensity was also significantly higher with the CE ToF MRA (*P*=0.01). This overall resulted in an increased CNR for the CE ToF MRA sequence, though there was no significant difference in the CNR for CE ToF MRA and standard ToF MRA (30.6±14.0 vs 28.9±16.0, *P*=0.08).

## Discussion

Our prospective study in patients with symptomatic lacunar stroke demonstrates that CE ToF MRA results in improved visualisation of the small cerebral perforating arteries compared with standard ToF MRA at 7T. The use of contrast was associated with improved visualisation and clarity of the lenticulostriate perforating arteries on our visual rating scale, and a greater visible length of the perforators. The improvement visualisation noted in our study, did not correspond with one previous study;^12^ this could be due to fewer subjects having pre-contrast ToF MRA were available for comparison.^12^ The greater visible length on the other hand, corresponds to findings in earlier studies.^12,19^ Use of contrast did not show a significant increase the number of perforators that were visible, similar to another study.^12^ On quantitative analysis, CE ToF MRA was associated with increased signal in the small perforating arteries although the increase in contrast to noise ratio was not significant. This could be explained by the increase in the background noise and brain tissue signal in the CE ToF MRA sequence. Another concern with the use of contrast is that it could increase venous contamination which would impair visualisation of the small perforating vessels.^12,20^ However, although venous contamination was slightly increased, this did not affect perforator artery visualisation. Therefore, our results suggest that contrast enhancement offers improvements in imaging of the small cerebral perforating arteries in this patient cohort.

Our study has a number of strengths. It was a prospective study with a predefined imaging protocol. All patients were imaged using an identical scanner and imaging characteristics, and standard ToF MRA and CE TOF MRA sequences were performed sequentially during the same imaging sessions and comparison of both sequences were made in the same participants at the same time. The two imaging sequences were assessed using a number of different analysis techniques which showed broadly consistent results. Visual rating was carried out by two experts blinded to each other’s results.

Our study, however, also has some limitations. The sample size was small, although by directly comparing results within the same patient between the two sequences, this was sufficient to show significant differences. We limited our study to the perforators arising from the MCA and ACA. The goal was to assess the LSAs, which most often originate from the proximal (M1) segment of the MCA^21^ as well as the proximal (A1) segment of the ACA.^22^ We used a narrow FOV with an acquisition time of approximately 10 minutes, making it difficult to include the whole Circle of Willis or other perforating arteries, such as the thalamoperforator arteries arising from the proximal posterior cerebral artery, posterior communicating artery and the tip of the basilar artery. In some cases, some LSAs could not be tracked beyond the area of coverage. Enlarging the FOV would invariably lead to increased acquisition time, prolonging the time a participant spends lying still in the scanner.^23^ A method of accelerating image acquisition without affecting image quality like compressed sensing^24,25^ has not been evaluated for CE ToF MRA; this is an area for further research. Thirdly, the use of contrast does have some potential side effects. Reports of nephrotoxicity in the development of a very rare condition of nephrogenic systemic fibrosis have been reported.^26^ For this reason, creatinine clearance was checked in all patients before imaging and any participants with impaired renal function were excluded.

## Conclusion

Currently, 7T MRI is largely used as a research technique, where it provides a unique opportunity to visualise the perforating arteries and their pathology. In such studies, the use of contrast enhancement improves visualisation of the LSAs and its use may aid in detecting pathology in cSVD, and in studying the effects of therapeutic interventions.

## Data Availability

Imaging data cannot be shared publicly because of ethical and legal restrictions to not compromise patient privacy. Imaging data are available from the Cambridge Service for Data Driven Discovery and access granted on application (contact via the corresponding author, C.N.O.) for researchers who meet the criteria for access to confidential data. All other relevant data are presented within the paper.

## Abbreviations

7T: 7 Tesla.
CE: contrast-enhanced.
cSVD: cerebral small vessel disease.
FOV: field-of-view.
LSAs: lenticulostriate arteries.
ToF MRA: time-of-flight magnetic resonance angiography

## Acknowledgments

We thank the Cambridge Central Research Ethics Committee for formal study approval, all the study participants and staff of the Wolfson Brain Imaging Centre, Cambridge.

## Author Contributions

All authors listed were responsible for aspects of study design, data collection and analysis and writing of manuscript, and meet criteria for authorship. Conceptualization and methodology, C.N.O., A.D.M., D.J.T. and H.S.M.; software and data curation, C.N.O., C.R. and C.T.R.; formal analysis, C.N.O. and M.E; rating scores, C.N.O and A.D.M.; writing—original draft preparation, C.N.O., and H.S.M.; writing— review and editing, C.N.O., A.D.M., M.E., C.R., P.B., D.J.T., C.R. and H.S.M.; visualisation, C.N.O., M.E. and H.S.M.; supervision, A.D.M., P.B., C.T.R. and H.S.M. All authors have read and agreed to the published version of the manuscript.

## Funding

The author(s) disclosed receipt of the following financial support for the research, authorship, and/or publication of this article: C.N.O. was funded by the Cambridge British Heart Foundation (BHF) Centre of Research Excellence (CRE, Centre Code: RE/18/1/34212) and a BHF project grant (PG/19/74/34670). H.S.M. was supported by an NIHR Senior Investigator award. Infrastructural support was provided by the Cambridge University Hospitals NIHR Biomedical Research Centre. This research was also supported by the NIHR Cambridge Biomedical Research Centre (BRC-1215-20014). The funders had no role in study design, data collection and analysis, decision to publish, or preparation of the manuscript.

## Institutional Review Board Statement

The study was conducted according to the guidelines of the Declaration of Helsinki and approved by the Institutional Review Board of East of England - Cambridge Central Research Ethics Committee (REC Ref: 19/EE/0219).

## Informed Consent Statement

Written informed consent was obtained from all participants in this study.

## Declaration of conflicting interests

The author(s) declared no potential conflicts of interest with respect to the research, authorship, and/or publication of this article.

Data cannot be shared publicly because of ethical and legal restrictions to not compromise patient privacy. Imaging data are available from the Cambridge Service for Data Driven Discovery and access granted on application (contact via the corresponding author, C.N.O.) for researchers who meet the criteria for access to confidential data. All other relevant data are presented within the paper.

## Notes

### Competing Interest Statement

The authors have declared that no competing interests exist.

### Clinical Trial

NCT04330222

### Author Declarations

This study was approved by the Institutional Review Board of East of England - Cambridge Central Research Ethics Committee (REC Ref: 19/EE/0219).

## References

1. Dichgans M, Leys D. Vascular Cognitive Impairment. Circ Res. 2017;120(3):573–591. doi:10.1161/CIRCRESAHA.116.308426

2. De Cocker LJ, Lindenholz A, Zwanenburg JJ, van der Kolk AG, Zwartbol M, Luijten PR, et al. Clinical vascular imaging in the brain at 7T. NeuroImage. 2018;168:452–458. doi:10.1016/j.neuroimage.2016.11.044

3. Zwanenburg JJM, Hendrikse J, Takahara T, Visser F, Luijten PR. MR angiography of the cerebral perforating arteries with magnetization prepared anatomical reference at 7 T: comparison with time-of-flight. J Magn Reson Imaging. 2008;28(6):1519–1526. doi:10.1002/jmri.21591

4. Cho ZH, Kang CK, Han JY, Kim SH, Kim KN, Hong SM, et al. Observation of the lenticulostriate arteries in the human brain in vivo using 7.0T MR angiography. Stroke. 2008;39(5):1604–1606. doi:10.1161/STROKEAHA.107.508002

5. Kang CK, Park CA, Park CW, Lee YB, Cho ZH, Kim YB. Lenticulostriate arteries in chronic stroke patients visualised by 7 T magnetic resonance angiography. Int J Stroke. 2010;5(5):374–380. doi:10.1111/j.1747-4949.2010.00464.x

6. Kang CK, Park CA, Lee H, Kim SH, Park CW, Kim YB, et al. Hypertension correlates with lenticulostriate arteries visualized by 7T magnetic resonance angiography. Hypertens Dallas Tex 1979. 2009;54(5):1050–1056. doi:10.1161/HYPERTENSIONAHA.109.140350

7. Riederer SJ, Stinson EG, Weavers PT. Technical Aspects of Contrast-enhanced MR Angiography: Current Status and New Applications. Magn Reson Med Sci. 2018;17(1):3–12. doi:10.2463/mrms.rev.2017-0053

8. Shin T. Principles of Magnetic Resonance Angiography Techniques. Investig Magn Reson Imaging. 2021;25(4):209–217. doi:10.13104/imri.2021.25.4.209

9. Kramer H, Runge VM, Morelli JN, Williams KD, Naul LG, Nikolaou K, et al. Magnetic resonance angiography of the carotid arteries: comparison of unenhanced and contrast enhanced techniques. Eur Radiol. 2011;21(8):1667–1676. doi:10.1007/s00330-011-2110-x

10. Zhou L, Xing P, Chen Y, Xu X, Shen J, Lu X. Carotid and vertebral artery stenosis evaluated by contrast-enhanced MR angiography in nasopharyngeal carcinoma patients after radiotherapy: a prospective cohort study. Br J Radiol. 2015;88(1050):20150175. doi:10.1259/bjr.20150175

11. Yang JJ, Hill MD, Morrish WF, Hudon ME, Barber PA, Demchuk AM, et al. Comparison of pre- and postcontrast 3D time-of-flight MR angiography for the evaluation of distal intracranial branch occlusions in acute ischemic stroke. AJNR Am J Neuroradiol. 2002;23(4):557–567

12. Harteveld AA, Cocker LJLD, Dieleman N, van der Kolk AG, Zwanenburg JJ, Robe PA, et al. High-Resolution Postcontrast Time-of-Flight MR Angiography of Intracranial Perforators at 7.0 Tesla. PLOS ONE. 2015;10(3):e0121051. doi:10.1371/journal.pone.0121051

13. Medicines.org.uk [Internet]. Gadovist 1.0mmol/ml solution for injection - Summary of Product Characteristics (SmPC) - (emc) [cited 2021 February 3]. Available from: https://www.medicines.org.uk/emc/product/2876/smpc

14. van der Kolk AG, Hendrikse J, Brundel M, Biessels GJ, Smit EJ, Visser F, et al. Multi-sequence whole-brain intracranial vessel wall imaging at 7.0 tesla. Eur Radiol. 2013;23(11):2996–3004. doi:10.1007/s00330-013-2905-z

15. Fan Z, Yang Q, Deng Z, Li Y, Bi X, Song S, et al. Whole-Brain Intracranial Vessel Wall Imaging at 3 Tesla Using Cerebrospinal Fluid–Attenuated T1-Weighted 3D Turbo Spin Echo. Magn Reson Med. 2017;77(3):1142–1150. doi:10.1002/mrm.26201

16. Li L, Chai JT, Biasiolli L, Robson MD, Choudhury RP, Handa AI, et al. Black-blood multicontrast imaging of carotid arteries with DANTE-prepared 2D and 3D MR imaging. Radiology. 2014;273(2):560–569. doi:10.1148/radiol.14131717

17. R Core Team [Internet]. The R Project for Statistical Computing [cited 2021 April 20]. Available from: https://www.r-project.org/

18. R Core Team [Internet]. Various Coefficients of Interrater Reliability and Agreement [cited 2021 November 6]. Available from: https://cran.r-project.org/web/packages/irr/irr.pdf

19. Umutlu L, Theysohn N, Maderwald S, Johst S, Lauenstein TC, Moenninghoff C, et al. 7 Tesla MPRAGE imaging of the intracranial arterial vasculature: nonenhanced versus contrast-enhanced. Acad Radiol. 2013;20(5):628–634. doi:10.1016/j.acra.2012.12.012

20. Hendrikse J, Zwanenburg JJ, Visser F, Takahara T, Luijten P. Noninvasive depiction of the lenticulostriate arteries with time-of-flight MR angiography at 7.0 T. Cerebrovasc Dis Basel Switz. 2008;26(6):624–629. doi:10.1159/000166838

21. Djulejić V, Marinković S, Maliković A, Jovanović I, Djordjević D, Cetković M, et al. Morphometric analysis, region of supply and microanatomy of the lenticulostriate arteries and their clinical significance. J Clin Neurosci. 2012;19(10):1416–1421. doi:10.1016/j.jocn.2011.10.025

22. Kang HS, Han MH, Kwon BJ, Kwon OK, Kim SH, Chang KH. Evaluation of the lenticulostriate arteries with rotational angiography and 3D reconstruction. AJNR Am J Neuroradiol. 2005;26(2):306–312

23. van der Kolk AG, Zwanenburg JJM, Brundel M, Biessels GJ, Visser F, Luijten PR, et al. Intracranial vessel wall imaging at 7.0-T MRI. Stroke. 2011;42(9):2478–2484. doi:10.1161/STROKEAHA.111.620443

24. Huang J, Wang L, Zhu Y. Compressed Sensing MRI Reconstruction with Multiple Sparsity Constraints on Radial Sampling. Math Probl Eng. 2019;2019:e3694604. doi:10.1155/2019/3694604

25. Park CA, Kang CK, Kim YB, Cho ZH. Advances in MR angiography with 7T MRI: From microvascular imaging to functional angiography. NeuroImage. 2018;168:269–278. doi:10.1016/j.neuroimage.2017.01.019

26. The Royal College of Radiologists [Internet]. Guidance on gadolinium-based contrast agent administration to adult patients [cited 2021 June 2]. Available from: https://www.rcr.ac.uk/publication/guidance-gadolinium-based-contrast-agent-administration-adult-patients

